# Social discrimination and stigma on the community of health professionals during the Covid-19 pandemic. An ethnographic approach

**DOI:** 10.1101/2021.10.28.21265608

**Authors:** Alexandros Argyriadis, Athina Patelarou, Vasiliki Kitsona, Alexandra Trivli, Evridiki Patelarou, Agathi Argyriadi

## Abstract

Severe Acute Respiratory Syndrome Coronavirus Type 2 (SARS-CoV-2) that caused the pandemic since March 2020, has affected among others, health professionals who work in covid-19 units by facing social discrimination. The aim of this study was to record the experiences of health professionals working in the first line of treatment of the pandemic, to analyse the effects of the pandemic on the interpersonal relationships of health professionals, and to ask about the stigma they faced during their work with people with covid-19.

This is a qualitative study with an ethnographic approach based on 160 semi-structured interviews with health professionals living and working in the Epirus Region, Greece. For the data collection we used semi-structured interviews, discussions and participatory observation. Specifically, the interviews were conducted on health professionals and more specifically doctors, nurses, rescuers, physiotherapists and administrative staff, working in covid-19 units at the University General Hospital of Ioannina (Reference hospital for Ioannina, in Epirus), which assists in the treatment of patients with covid-19, and in the branch of the rescue department of Ioannina.

The data were analysed in four thematic units based on their common characteristics: a) emotions and experiences of health professionals, b) interpersonal relations of health professionals, c) social exclusion and discrimination, and d) health professionals as patients. The results showed that the main emotions that health professionals experienced when they were moved to covid-19 clinics were fear, anxiety, distress, anger and insecurity. These feelings worsened when their family environment treated them with fear and hesitancy. Their social environment tended to avoid them, leading to a state of self-isolation. To conclude, health professionals faced discriminating behaviors and stigma both from their families and social environment, and from other health professionals. The government struggled to handle the situation in keeping a balance between both the security and well-being of health professionals as it was not prepared for a pandemic like this.

## Introduction

With the spread of the coronavirus, concerns prevailed regarding the effects of the pandemic on the health and economy of the population (Harapan et al., 2020) while at the same time there were many negative effects of the pandemic on health professionals (Bhanot et al., 2021; Murphy et al., 2020). One of the major concerns that is gradually emerging from recent research is the strong bias against health professionals working in units with covid-19 patients (Bhanot et al., 2021). It has been noted that the white uniform became a symbol of infection during the Pandemic Crisis (Singh and Subedi, 2020).

The discrimination that the above employees have received concerns the refusal of dialogue and cooperation of a portion of the population with employees in covid-19 spaces, the refusal to enter restaurants and dissatisfaction about having to stay at home. This is all despite the fact that the employees in the field of health care work take all necessary precautions and are specially trained (Brooks et al., 2017). In several countries, such as the United States, India, and Australia, reports of attacks, threats, violence, and even persecution have been reported in newsletters (Singh and Subedi, 2020; Xiong and Peng, 2020). The above reaction could be interpreted as an expression of society’s fear of those who come into contact with the infectious disease covid-19 and a deeper fear of being infected.

High levels of transmission in the community, in healthcare settings, and in family and close friends also enhance the sense of danger and threat (Patelarou et al., 2021). All the above have led the majority of the population to turn to negative behaviours, manifested by avoiding intercourse by isolating people who are likely to be infected (e.g., people traveling abroad, working in health facilities or who have come into contact with a patient). Emotions recorded by relatives and other citizens who came in contact with health professionals were mainly feelings of rejection, fear, and categorisation (Duan et al., 2020). Recent research, which is extremely limited on this issue due to the fact that this outbreak is quite recent, has started to record the experience of health professionals during the pandemic, although there is no similar research in Greece and Cyprus (Eftekhar et al., 2020; Gordon et al., 2021; Montgomery et al., 2021; Smyrnakis et al., 2021). It is characteristic that such opinions were expressed in the daily press during the pandemic period about health professionals that were forced to separate from their families and live in hotels, thus preventing further spread of the virus. Such a situation was applied in Cyprus with adverse effects mainly on the psychology of professionals (Taylor et al., 2020). However, the appearance of social stigma towards health professionals seems to be significantly influenced by the country in which it is studied. Thus, in many Asian countries (China, Hong Kong, India and Nepal) but also in the United States, there is a strong element of stigma towards health professionals. In contrast, in the United Kingdom, a small proportion of the population treats staff associated with and treating covid-19 patients with racism (Argyriadis, 2020; Bagcchi, 2020).

It is interesting that there is social stigma among health professionals as well. In particular, staff who are not involved in caring for covid-19 patients often have a negative attitude towards health professionals who are. In such cases, phenomena such as refusal of social contact e.g., not staying in the same place for eating with staff who are involved covid-19 patients, have been observed. Moreover, social racism is observed at the neighbourhood level, since phenomena of dissatisfaction have been recorded regarding staying in the same apartment building with health staff who work with patients with covid-19 (Singh and Subedi, 2020).

Similar phenomena have been reported in other pandemics in which employees in institutions and clinics treating patients with communicable diseases experienced higher levels of stigma compared to those working in other clinics (Adja et al., 2020). Due to the significant lack of similar studies, it was deemed necessary to study this issue in order to highlight the experiences of health professionals and this study should be a trigger for further research on the issue.

### Aim of the study

The aim of this study was to record and analyse the expression of social exclusion in health professionals who directly care for patients with covid-19. In particular, we aim to understand the main behaviours faced by health professionals from the social environment when people hear that they are at the forefront of the covid-19 virus, the psychological effects of social exclusion on health professionals, and the feelings of those involved.

The main questions that arose from the literature review were:

- What are the main behaviours faced by health professionals from the social environment when people hear that they are at the forefront of the Sars-CoV-2 virus?
- What are the psychological consequences of social exclusion behaviours for health professionals?
- What are the effects of the pandemic on the social activity of health professionals?
- How do health professionals feel and behave as patients with covid-19?

## Method

The method that was deemed most appropriate for the design of this study was the qualitative one and more specifically the ethnographic because the research team wanted to pursue an in-depth study of human beliefs, attitudes, perceptions, ideas and experiences. Regarding the data collection we used participant observation, semi-structured interviews and informal discussions. The design of a semi-structured interview guide was constructed for the needs of the current study, which was piloted before its use. The interview guide was structured in five thematic units. In the first thematic unit, the personal data of the participants were recorded, in the second thematic unit, emphasis was placed on the mental health of health professionals, during their work in covid-19 clinics. In the third, the impact of the pandemic on the interpersonal relationships of health professionals, the fourth unit examines social exclusion during the pandemic and the fifth and final unit focuses on those health professionals who became ill with the virus and its effects on their mental health, daily life and interpersonal relationships.

### Research process

Due to the lockdown that was imposed by the Greek government since November 2020, but also for security reasons, the research team decided to conduct interviews via Skype, Zoom, WebEx and other means that could offer communication with audio and video. The first step for the implementation of the project was the pilot study of the interview guide by 16 pilot interviews (10% of the total sample), in which the questions were tested. The pilot test resulted in changes and corrections mainly at the verbal level so that the questions were more understandable. The pilot interviews were successful, so the guide was applied to the remaining interviews. Prior to the interview, a telephone contact was held in which the reasons for the interview were explained, the necessary information was provided and an e-mail with the consent form was sent. After the oral and written consent were given, the interview was conducted and videotaped for the purposes of analysis. At the end of the interviews, answers were transcribed and categorised according to the thematic unit to which they belonged. The main findings were evaluated and a thematic analysis was performed in order to lead the team to the final conclusions.

### Sample - Context

The study focused on the Region of Epirus, Greece and specifically the Prefecture of Ioannina during the months of January and June 2021. The specific geographical location was chosen due to the reduced research activity in the area and the outbreak of violence against health professionals, as presented in news bulletins. Specifically, the interviews were conducted on medical staff and more specifically doctors, nurses, rescuers, physiotherapists and administrative staff, working in covid-19 units at the University General Hospital of Ioannina (Reference hospital for Ioannina, in Epirus), which assists in the treatment of patients with covid-19, and in the branch of the rescue department of Ioannina. A total of 160 interviews were conducted. The mean age of the sample was 47.1 ± 6.9 years. The method of snowball sampling was followed in the hopes of reaching out to other people who have felt stigmatised, either themselves or those close to them. By snowball sampling, individuals who already had agreed to participate in the study could recommend others to share their experience with the research team. The basic criterion for selecting a sample was that they needed to work in units that treat patients who are positive for the Sars-CoV-2 virus and to have direct contact with patients. At the same time three members of the research team were working at the General Hospital of Ioannina and the emergency – rescue center in order to proceed with the participatory observation. These members were keeping field notes and recordings and they conducted the informal discussions and interviews.

## Results

The results of the research were categorised into four thematic sections: a) the study of the feelings of health professionals from their experience, and the behaviour of the social environment, b) their interpersonal relationships within clinical settings, the social exclusion they received from the wider social context, and from their experience as patients.

140 of the 160 respondents were over 40 years old and the oldest was 63 years old. 69% of the respondents were women and the majority were married and living with their families. Finally, 69% of the sample were nurses, 12% doctors, 2% physiotherapists, 5% rescuers and 12% administrators. It is important to note that only 24% of them had been or were currently infected by the virus.

### A. Experiences and emotions of health professionals

The first topic studied the emotions and experiences that health professionals had developed from their work in an environment with a high viral load, and also the changes brought about by the internal transfers of covid-19 clinic staff to their mental health and work experience. In the questions that were related to the emotions that overwhelm the participants the results were mainly negative (158 participants) with fear prevailing in almost all respondents. The other emotions recorded were frustration, sadness, anxiety, despair, fatigue, anger and insecurity. Other negative emotions recorded were panic and guilt while others said they were more closed and wary of those around them.

Typical excerpts from the interviews are:

> *“I feel intense anxiety, fear, mental fatigue and bitterness towards a set of people who think that we are not doing something great”* (P1).
>
> *“Initially I felt stress, I had a feeling of fatigue and exhaustion. Negative thoughts about not being able” (P23)*.
>
> *“I panicked and I was scared for my child, my husband and parents, so I felt terrible anxiety”* (P59).
>
> *“I also felt guilty, and I will explain to you why. Because on the one hand I was afraid for myself and my family, but on the other, I did not want to go to be with the patients”* (P152).

But in addition to the negative emotions, some respondents felt satisfaction and joy that they were at the forefront of tackling the pandemic. Specifically, the excerpts related to positive emotions are:

> *“I feel sad, I feel that I have lost things all this time, but the feeling that prevails is that of joy because of what I have been able to offer. Emotions are mixed “* (P70).
>
> *“I found out who are the people close to me and on whom I can count”* (P10).

When the participants were asked what has changed in their lives and their work after this experience, they said that they are more suspicious, more careful, and that the workload has increased, since in addition to the serious condition of the patients, they must take care to observe and adapt to changes. The changes are related to the continuous evolution of measures adopted to protect and prevent the spread of the virus. Typical excerpts recorded were:

> *“I pay more attention than usual. I change my gloves and mask more often”* (P29).
>
> *“I am exposed to something unknown… an unpleasant feeling of inadequacy”* (P31).
>
> *“The government constantly changes all protocols. We have to adjust to the new conditions and make too many changes in our everyday routine”* (P12).

Finally, asking about the mental health effects of the pandemic, the main views expressed focused mainly on future predictions. Respondents believe that isolation due to the lockdown will have a negative impact on the population. Some people also see an increase in incidents of social racism. This is also confirmed by characteristic excerpts such as:

> *“I think there is strong suspicion and fear around. People are afraid of getting sick, they are very careful with whom they come into contact, and they feel intense anxiety and discomfort with the whole situation”* (P127).
>
> *“I noticed that there is a strong phenomenon of social racism and a mood of the people to comment, gossip and find out who is sick and who is not, and who is not”* (P12).
>
> *“I think that there is an outbreak of stigma on those who are sick or who are in the environment of the sick”* (P83).

However, there is also a portion of respondents who believe that the pandemic will also leave positive elements. The most optimistic views are related to the fact that some good habits about hygiene will be adopted. They also believe that after this challenging experience in difficult conditions, that most people will be more resilient and will be able to manage their lives more optimistically. Finally, they believe that all of this will make the health system stronger and that the professionals will improve their skills. Indicative answers are listed below.

> *“We will come out more armored and stronger! Both our emotional endurance and our relationships have been tested. I already feel stronger”* (P55).
>
> *“Society has realised that we are useful, and this means a lot to us”* (P105).

### B. Interpersonal relationships of health professionals

In the second thematic unit, the questions focused on the interpersonal relationships between health professionals and their families, and also among health professionals after they had moved to clinics and departments related to the care of covid-19 patients. Most health professionals (103 participants) said that their family was scared by the announcement of their transfer. The second dominant emotion recorded was anxiety (122 participants), while feelings of intense anxiety were also noted, they avoided meeting them, feeling frustrated, sad and prejudiced (94 participants). Of course, there were families who did not react differently, but the health professionals isolated themselves to protect them. The main excerpts highlighting the emotions that were expressed are the following:

> *“Look, at the beginning, there was intense fear; they avoided meeting us. They were terrified”* (P22).
>
> *“I saw in their eyes sadness and disappointment… they also treated me with fear and prejudice… then they got used to it”* (P36).

Negative behaviors were also observed by colleagues of health professionals (39 records). Either colleagues who worked in other departments not related to covid-19 patient care, or hospital staff. The exact opposite picture, however, was presented for colleagues who belonged to the same department and the same clinic. They described the climate as positive, with support, understanding and solidarity. The main negative emotions that were expressed were the avoidance of socialising, caution and even alienation in their relationships, mainly from the administrative staff.

> *“Basically, I feel stigmatised and this makes me very sad, because I did not expect it from my colleagues”* (P13).
>
> *“Some people avoid you. They asked us not to stay together because we work on a covid-19 clinic. They did not want to eat with us at the same table so as not to be too close. They did not want to have any kind of contact”* (P62).
>
> *“We have been targeted by our colleagues”* (P70).

In terms of relationships, differentiations in the care and treatment of patients with covid-19 compared to other patients were also assessed. The nature of the disease (it is something new and unknown), the nature of the hospitalisation, and the management by the media have terrified patients, they seek communication and encouragement from the nursing staff as they are the only ones who associate with them, not their families. The hospitalisation of these patients, characterised by the absence of attendants, results in them feeling terribly lonely for the entire period they are hospitalised and seeking psychological support from health professionals.

> *“They seek communication with the doctor. They need to be encouraged that they will get well, and they try to keep longer discussion”* (P11).
>
> *“They are terrified, I would say isolated patients are too afraid. Nurses are the means by which they communicate with the outside world”* (P52).
>
> *“Covid patients have problems with their survival. They do not know if they will come out alive and intact in relation to other patients… they are more grateful patients”* (P5).
>
> *“These people need more support, psychological support”* (P7).

### C. Social exclusion and discrimination

The third topic focused on the social discrimination of the local community against health professionals, which was one of the main topics of this study. The participants’ responses indicate social discrimination, alienation, specific negative characterisations, defensive attitudes and other similar attitudes and behaviours. In particular, most participants (136) stated that the behaviours they have observed and accepted from their social environment, were the avoidance and marginalisation that led to the feeling of not having a social life. Behaviours of self-isolation from the side of health professionals were due to the fact that either they did not want to embarrass those around them, or they did not want to be confronted with bad behaviours and feel marginalised by their friends. The majority of respondents isolated themselves and avoided social activity.

> *“Any social activity I had in the past has been eliminated. I avoid social activities, so as not to embarrass others because of my work. Mental and physical fatigue leads me not to have the mood for socialising” (P17)*.
>
> *“We are afraid to come in contact with other people because since we work in such an environment, it is dangerous to be at the same place with others. I feel that I am excluded from my social network and I am trying to avoid unpleasant attitudes” (P23)*.
>
> *“They try to keep a distance from me” (P35)*.
>
> *“I do not want to suggest a dinner or anything because I feel that I might put them in a difficult position. I constantly find excuses” (P6)*.
>
> *“I used to hide where I work, I was really scared. Because I know that she will get stuck, that they will see me somehow, that they will be scared, that I will cause stress to them” (P78)*.
>
> *“I have reduced my social activities to a minimum! I would say that I am not in the mood to do anything and seeing the behaviour of the people here in the neighbourhood, who are afraid of me, who avoid me” (P11)*.

Most of the health professionals experienced negative behaviours from their social environment, by friends, acquaintances, tenants of apartment buildings, and also behaviours in neighbouring stores such as mini markets and bakeries and gave several examples of these behaviours. In the behaviours recorded, most said that those around them avoided contact with them, while a large portion of the respondents also stated that they felt alienated from their social environment and that they did not want to contact participants in activities that until then they had done together. The most representative answers are the following:

> *“The social environment, such as my neighbours avoid any personal contact with me and I have also heard negative comments from the opposite sidewalk. When I meet people that I know in front of a cash register, they leave miles away from me or wait for me to leave and then proceed to their shopping. My friends are worried or completely avoid socialising with me. They communicate only by phone and show their anxiety” (P1)*.
>
> *“People disappear when they hear that I work at a covid-19 clinic” (P52)*.
>
> *“I was climbing the stairs to my apartment and I met a tenant of another apartment staying still as soon as she heard me coming down. She was waiting for me to enter the door of my house and then she came down. I felt very bad, very disadvantaged; the lady treated me with such racism and with such disrespect. Racism in general, is too intense in the wider social context” (P7)*.
>
> *“They avoid me and at some point even my neighbour said to me: okay now, you are also there at the hospital, you are in contact and yes we are afraid” (P92)*.
>
> *“My friends don’t see us anymore, they don’t even want to go out for a walk together” (P11)*.
>
> *“I was talking to a friend of mine and I was describing the possibility of going to Konitsa on Saturday to meet him. He answered: Look, don’t come and bring me the virus” (P16)*.

### D. Health professionals as patients

24% of the sample said they tested positive for covid-19 and became ill. This result led the research team to design another thematic section in order to study how health professionals felt as patients and what behaviours did they face. The results of the study showed that all patients were terrified, anxious and panicked with the announcement of the disease, while loneliness was the key emotion. Fear was also a key emotion, both for the outcome of their health and for the health of their loved ones. At the same time the behaviour of some of their colleagues was empathetic, warm and supportive but from some others who work in other wards it was quite uncommunicative.

> *“I felt dissatisfaction, fear, insecurity, anxiety about the outcome of the disease” (P1)*.
>
> *“Well, I feel very lonely… A lot of anxiety about my life, a lot of stress; I felt like I was losing the earth under my feet” (P9)*.
>
> *“Fear and anxiety, more about my children and my wife” (P15)*.

Their social environment also expressed a series of negative emotions as some were: *“completely lost” (P93)*, and experienced suspicion and fear as well as disrespect *“they looked at me like I had leprosy” were the main emotions*.

> *“There was intense suspicion and fear; Ioannina is a closed society. I felt like I was seeing a wall between me and the others, that they were avoiding me” (P107)*.
>
> *“They looked at us as if we had leprosy” (P151)*.

These behaviors continued even after the respondents recovered and had a negative PCR result since their social environment continued to avoid them and to be suspicious.

> *“Those around me treated me with hesitation, with fear, despite the fact that now my PCR test is negative negative” (P1)*.
>
> *“People did not believe that I have a negative result” (P9)*.
>
> *“I was treated with suspicion and fear. Indicatively, when I returned to the café of the hospital where I work, the barista who saw me was looking to find where his mask was as he was not wearing a mask. A couple of people who were inside, sitting, got up from their chairs and started to leave” (P10)*.

Similar behaviours were observed by the professional environment.

> *“Some colleagues of mine withdrew every collaboration with me, even though I was now negative on the test” (P1)*.

## Results from the observation

The field observation showed that men were more aggressive or avoided cooperating with people who were in the front line of covid-19. There have been cases where social discrimination has focused mainly on the body, which has suddenly become morbid, pathological, and therefore dangerous. They were therefore in favor of limiting those who came in contact with positive cases. The expression of social discrimination was not limited to oral speech but mainly to non-verbal expressions of disgust, fear and avoidance. In addition to gender differences, there were also social differences depending on the educational level of the participants in the research with people who had postgraduate and doctoral studies being more friendly and showing understanding. Age was another interesting factor which showed that younger ones accepted more easily the diversity compared to the older ones who were often absolute or more fearful. Finally, it is important to note that the human body could be symbolised in multiple ways depending on the context in which it was examined. Other times certain people were described as heroes, other times as infectious and dangerous and other as ordinary workers.

## Discussion

All participants stated that the announcement of their transfer to clinics or departments directly related to covid-19 patients led to the manifestation of negative emotions such as fear (the most common response given), anxiety, distress, anger and insecurity. Many of them stated that they were forced to move to the new department and that it felt like being punished. These feelings are also partly encountered in the literature, where a large portion of those who were transferred to a ward related to covid-19 patients were anxious and frightened (Sun et al., 2020). In particular, one study reported that all employees were negative and frightened during their transfer to a new ward, while their fear peaked during the first days of work in a negative pressure ward with covid-19 patients. Another study stated that it was their duty and joy to take part in the fight against the virus and that they would do their best to deal with it as soon as possible (Liu et al., 2020). Emotions that are justified due to the high contagion of the virus, but also the fear of the unknown, which most of the times causes panic. Several participants stated that this feeling of fear and insecurity was due to their lack of information and lack of training before taking up their new duties. Also, the lack of equipment in addition to limited staff for these departments also led to the feeling of panic. It is important to note that a key factor in denial was the participants’ fear of transmitting the virus to their families and loved ones. Proper education and information leads to a sense of self-confidence and that they are properly trained to deal with viruses. A study conducted in the USA in 2004 showed that 70% of employees do not want to work in an environment where instruction and training are insufficient, results that are consistent with those of our study (Person et al., 2004). In many countries, such as Sudan, the families of health professionals exert further psychological and physical pressure (Çiftci et al., 2021). The most accurate information, the right education and the lack of misinformation, but even the psychological support according to the literature has been shown to reduce these negative emotions and the supportive environment can help deal with psychological fatigue resulting in more efficient work (Liu et al., 2020).

This change in the workplace also brought about changes in work experience, as everyone stated that they became more suspicious and cautious, while the workload increased significantly. These data agree with the literature where the same feelings of dissatisfaction have been recorded, as in this study which states that health professionals are psychologically and physically tired from time-consuming protocols and the increased workload (Sun et al., 2020).

The impact of the pandemic on interpersonal relationships is also significant. The literature has studied and continues to study the effects of pandemic and social exclusion on interpersonal relationships. One big issue, however, is the impact on the relationship between health professionals and those around them. History has shown that in epidemics and pandemics, people are divided, and there are those who understand the difficult work of health professionals, but also those whose sense of survival leads to the manifestation of behaviours of marginalisation, alienation and even racism towards the medical staff. It has been seen in studies from previous pandemics or epidemics, where people are divided into three categories; those who believe in the existence of the virus and perceive the work of health professionals and those who do not believe or who marginalise them. Our results are consistent with two large studies conducted in Sierra Leone and the Congo in 2015, which showed that all nurses working with Ebola-positive patients were marginalised (Kimball et al., 2020; McMahon et al., 2016). The majority of respondents said that their family expressed negative feelings in the announcement that they will now work in the first line of care for patients with covid-19, as almost all said that their family was scared and anxious about this transfer and there were cases of marginalisation in the same house (separate sleeping for couples, separate celebrations and distancing). The treatment of friends, acquaintances and neighbours was also negative since the avoidance of contact, alienation and behaviours of social racism were recorded by the respondents. According to the literature, these behaviours are common, since cases of violence, exclusion from restaurants and even eviction from their own home due to their work in a hospital have been reported (Kackin et al., 2021).

Health professionals also expressed differences in the treatment of covid-19 patients compared to other patients. What everyone has said is that patients’ psychology is very different. Patients feel alone, isolated. They have no relatives present, there is no direct contact with the staff, and the length of hospital stay is long, which they spend alone. The fact that it is an unknown disease, about which much is said in the media and that little is known about its development leads to concern and anxiety about their health. They seek encouragement and communication from nurses and psychological support. These findings are consistent with the literature, since the feeling of loneliness is predominant in patients and is due to the way of treatment and the lack of accompaniment (Gao et al., 2020). The implementation of lockdown has led to the reduction and minimisation of human interactions, social contact and activities. However, the social activity of health professionals has been eliminated not only due to the lockdown but also due to the marginalisation they experience from their social environment. Behaviours of racism and distancing have positively fuelled social isolation and in order not to embarrass anyone or to feel bad about the behaviours they are confronted with, they have isolated themselves and abandoned their social life. This behaviour is confirmed by the existing literature as it seems that the behaviour of withdrawal from social life due to mental fatigue and behaviours that prevail in the social environment is typical, with the result that health professionals are further burdened by social isolation (Ardebili et al., 2020). Another fact that was highlighted by the literature review and confirmed by our study is that there were negative behaviors from colleagues who do not work in covid-19 related departments, who come in direct contact with covid-19 patients, or by administrators, as shown by a study conducted through telephone interviews in Iran. This research showed the same results as ours, where colleagues of professionals working with covid-19 patients, avoided them by offering more stress in an already difficult psychological situation (Galehdar et al., 2020).

Finally, the treatment of health professionals as patients with the virus was highlighted, where the picture was full of negative emotions received both from relatives who helped them during their illness and most of their friends, even after they had returned healthily to work for a long period of time. This result is consistent with a case study published in 2020 in which a physician who had symptoms similar to those of covid-19, despite undergoing a molecular test which was negative and after a period of quarantine, on her return she was ridiculed and resented by her social environment. Facts that show that fear of disease can lead to extreme behaviors (Grover et al., 2020). Health personnel throughout the pandemic are making superhuman efforts against a new threat. The quantity of staff is minimal, and the needs are numerous. In order for the staff to be in the front line, they must have all the supplies (equipment, training) but also remain mentally and physically healthy. Mental health is vital and is an issue that is evident in the present study. Health staff must be psychologically supported, and this must be a concern of every hospital institution but also of the state. Psychological assistance should be provided to all health professionals, especially to those working in covid-19 departments. Also, an issue that has emerged is the need to defend the labour rights of nurses who come into daily and long contact with covid-19 patients. Everyone stated that they would like the State to show its support in practice, including their work in the “heavy - unhealthy professions”, as stated in the official announcement of the Panhellenic Nurses Association. In addition, they seek gratitude, not only on the level of “applause”, but that the State be able to show it at the level of pay in these difficult days we are experiencing (O’Brien, 2003).

## Conclusions

From the present study, it emerged that in the first thematic unit regarding the emotions and experiences of health professionals, the main emotions experienced by the participants were fear, anxiety, fatigue, agony, frustration, sadness, anger, insecurity and panic. These feelings started mainly when they heard about their transfer to their new work environment and because of insufficient information regarding the sudden changes in their work environment.

In the second unit on interpersonal relations of health professionals, a strong picture of alienation from the social environment was observed with friends, neighbours and acquaintances marginalising and avoiding any contact with health professionals. However, key issues arose which concern the lack of training of health professionals and the need for better information, as well as the need for recognition by the State. They ask for this recognition to be “translated” into payroll and through joining the category of heavy and unhealthy professions. The most important issue however, was that there was social discrimination and inequality among health professionals. In the third section of the study regarding social exclusion and discrimination, the phenomenon of self-isolation was also strong as a defence against “racist” behaviours from those around them. It was particularly traumatic to impose self-isolation on health professionals and force them to stay in hotels away from their families.

Finally, regarding health professionals as patients, they seemed to be emotionally burdened because they were faced with the fear, stress and anxiety of their loved ones. They gained experienced and knowledge of dealing with the virus and faced the fear of death and the social stigma of others.

Finally, they were also adversely impacted by exclusionary behaviours from their colleagues, either health or administrative.

## Data Availability

All data produced in the present study are available upon reasonable request to the authors

